# Factors Associated With Chronic Arthropathy And Rheumatological Diseases After Chikungunya Infection in Colombia: A Case-Control Study

**DOI:** 10.1101/2024.05.23.24307784

**Authors:** Guillermo Alejandro Ramírez Luna, Fabian Méndez Paz

## Abstract

**Objective:** To identify the factors associated with the development of chronic arthropathy and rheumatological conditions in individuals who presented with clinical chikungunya infection.

**Methods:** A case-control study was carried out among individuals with a clinical diagnosis of chikungunya treated at a Colombian Health Provider Institution between 2014 and 2015. Two logistic regression models were constructed: one excluding the variables with more than 50% of the missing data, and another with all the variables but with the imputed data.

**Results:** In total 133 cases and 216 controls were obtained (349 individuals). In both models, female gender was a risk factor (OR: 2.27, CI95: 1.03 - 4.97, p = 0.04, and OR: 2.37, CI95: 1.09 - 5.17, p = 0.029, respectively), while acute involvement of the wrists was a protective factor (OR: 0.44, CI95: 0.20 - 0.95, p < 0.037; and OR: 0.29, CI95: 0.13 - 0.65, p = 0.003 respectively). Arthralgia duration was a risk factor in the imputed data model (OR: 1.03, CI95: 1.00 - 1.06, p = 0.02).

**Conclusion:** Female gender and the duration of joint symptoms in the acute phase were the main risk factors for the development of chronic chikungunya arthropathy or rheumatological pathology. Wrist involvement was a protective factor. Only rheumatoid arthritis was found to be a rheumatological disease associated with chikungunya infections. Finally, this study suggests that the interaction between usual cardiovascular factors could enhance the probability of developing chronic symptoms.

## Introduction

Acute chikungunya disease (CHKGV) is a viral infection that manifests as a critical febrile illness accompanied by exanthema and intense generalized arthralgia or arthritis which becomes chronic in some individuals. (1,2) The term “Chikungunya” comes from Mozambique, Africa, and means “to be inclined” alluding to the posture adopted by the intensity of arthralgia. The chikungunya virus is an alphavirus belonging to the Togaviridae family and is mainly transmitted by Aedes albopictus and Aegypti mosquitoes. Since 2013, the chikungunya virus has been endemic in the Americas, including Colombia, where the epidemic occurred from 2014 to 2015 in almost the entire national territory located 2.200 below sea level, with more than 123 municipalities affected. The most frequent complication is chronic chikungunya arthropathy, which consists of severe polyarticular pain that persists for more than 3 months and affects 30% of infected individuals (3–7). The appearance of rheumatological conditions such as rheumatoid arthritis, inflammatory spondyloarthropathy, systemic lupus erythematosus, and Sjögren syndrome after infection has also been described (7–12). On the other hand, in rhe general population there are associations between rheumatological diseases such as a higher probability of presenting with rheumatoid arthritis if there is a family history of systemic lupus erythematosus, Sjögren’s syndrome, or polymyalgia rheumatica (13) as well as frequent joint involvement, some immunological markers, the response to immunosuppressive drugs, and in general the highest prevalence in women. It seems that those infected with CHKGV are at risk of developing some of them, but in addition, joint symptoms due to chikungunya can be indistinguishable from early rheumatoid arthritis to the point that rheumatoid arthritis classification criteria have been used for its diagnosis (8,12,14–18).

Globally, factors associated with the development of chronic chikungunya arthropathy have been described, including advanced age, female sex, history of osteoarthritis, the severity of clinical presentation, elevated levels of C-reactive protein, elevated levels of IgG, persistence of virus RNA detection beyond 7 days after the onset of symptoms, and absenteeism from work, among others (7,17–26).

In Colombia, studies have evaluated the factors associated with the development of chronic arthropathy only in the state of Tolima and the state of Atlantic. However, the results have been dissimilar (15,21). Valle del Cauca is one of the main states of the country and is one of the most affected by the Chikungunya epidemic; however, there is a lack of studies in this regard. It is highly probable that new epidemic outbreaks will occur, therefore it was decided to carry out a case-control analysis in a population of such a state to identify the factors associated with chronic arthropathy and/or rheumatological conditions in individuals who presented clinical infection by Chikungunya.

## Subjects and methods

### Study design

A case-control study was conducted among individuals with a clinical diagnosis of Chikungunya treated at a Health Provider Institution (HPI) between 2014 and 2015. The HPI are entities of a private, public, or mixed nature authorized to provide the health services contemplated in the Colombian Obligatory Health Plan. The HPI of this study is located in the state of Valle del Cauca, and serves approximately 6,200 individuals per year, mainly belonging to socioeconomic strata 3, 4, and 5 (there are 6 strata in Colombia and they are categorized from lowest to highest as socioeconomic conditions improve) and with 95% affiliation to the contributory regime (salaried individuals and their family nucleus). Valle del Cauca, Colombia, is the second most developed department in the country and had a population of 4,600,000 inhabitants until 2015

### Definitions

A case was defined as any individual who developed any of the following conditions after infection: chronic inflammatory arthropathy, systemic lupus erythematosus, Sjögren syndrome, seropositive or seronegative rheumatoid arthritis, rheumatic polymyalgia, or inflammatory spondyloarthropathy. Chronic inflammatory arthropathy is defined as the persistence or presence of inflammatory arthralgia(s) after 3 months of the acute phase of CHKNV infection, as long as other causes of chronic inflammatory arthropathy, such as the pathologies previously mentioned, have been discarded (see definitions in the Supplemental Digital Content 1).

The controls were the individuals who presented with clinical chikungunya infection but did not develop any of the mentioned pathologies.

It was decided not to match for any factor considering that, since it is a case-control study, this procedure can bias the measure of association towards the null value when the exposure variable is related to the confounder (36)

### Sample size calculation

Using the Fleiss method (27) a sample size of 135 individuals was estimated for an alpha error of 5%, a power of 80%, a prevalence of exposure “age over 45 years” in controls of 59%, a ratio of 2:1 between controls and cases, and an estimated data loss of 20% out of an expected OR of 3.9 (7).

Finally, the cumulative incidence of different rheumatological diseases evaluated in this study was explored in the population that made up the sampling frame using the ICD-10 codes described in Table A1 (Supplemental Digital Content 2). The frequency of chronic arthropathy due to chikungunya was not performed on the sample frame because there was no specific ICD 10 for this condition. However, they were calculated from the total reviewed records. In the case of polymyalgia rheumatica, the calculations were performed on patients aged 50 years or older.

### Data collection

The HPI of the study provided the medical records of the sample frame, which was selected using ICD-10 A920. The ICD-10 codes described in Table A1 (Supplemental Digital Content 2) were used to search for cases. The medical records of the cases and controls were selected using simple random sampling, which were previously coded to provide anonymity, however before taking the sample, one of the author had access to all the information for the purposes of coding the clinical histories. For the definition of chikungunya infection, the clinical criteria of the attending physician were used as long as the presence of fever and arthralgia was recorded in history. The period in which the sampling frame was extracted was from July 1, 2014, when the first case of chikungunya was recorded in Colombia, to December 31, 2015, which corresponded to the end of the epidemic. The data access was obtained in august 2019.

Taking into account the difficulties in Colombia to obtain confirmatory laboratory tests in most suspected cases of Chikungunya, it was decided to use the clinical criteria of the attending physician, as long as the presence of fever and arthralgias, appealing to the classification criteria of the WHO (37). The period selected to collect the sample was the one with the highest number of cases recorded in Colombia to ensure a higher positive predictive value for the clinical definition of chikungunya.

The inclusion criteria were individuals aged 16 years or older with a clinical diagnosis of Chikungunya infection.

Individuals diagnosed with the rheumatic disease before chikungunya infection and those receiving immunosuppressants or any drug used to prevent transplant rejection, control rheumatic diseases, or any curative treatment of cancer including chemotherapy were excluded.

Sociodemographic factors, the pathological history of individuals, and clinical factors associated with infection were explored. Information on socioeconomic status was unavailable.

The study was approved by the Ethics Committee of the Faculty of Health of the Universidad del Valle CIREH and the Ethics Committee of the HPI from which the study subjects came (approved record 007-020), which waived the requirement for informed consent.

### Statistic analysis

Normality was evaluated using the Shapiro-Wilk test. Means and standard deviations were used as measures of central tendency and dispersion, respectively, and Student’s t-test was used for comparison between groups. Medians and interquartile ranges were used for variables that did not meet the normality criteria, and the Wilcoxon and Kruskall-Wallis tests were used for comparison between groups. For the “weight” variable the mode was used. Logistic regression and 2 × 2 tables were used to obtain raw ORs with their respective 95% confidence intervals. To facilitate the analysis, age was dichotomized into “greater than or equal to, and under 40 years of age”, body mass index normal/low was grouped into “low” and overweight and more into “high”, and educational levels corresponding to primary school, preschool, and no education were grouped into “low”, and the rest of the levels as “high”.

Co-linearity was evaluated using the Pearson’s test. For the adjusted analysis, three logistic regression models for estimate OR were made using the stepwise method: one with the sociodemographic variables and background of the individual, another with the clinical variables of the infection, and the last one with the laboratory variables. Subsequently, a model was made with the variables of the 3 models that had a statistical significance of less than 0.2.

Given that there was a decrease in the sample size of almost 90% with this model, two models were created: one excluding the variables with more than 50% of the missing data and another with all the variables but with the imputed data using the multiple imputation method.

The Goodness of fit was estimated using the Hosmer test, Pearson’s residual and deviance analysis, and the Bayesian information criterion (BIC).

Two sensitivity analyses were performed to evaluate the misclassification bias for chikungunya infection using the diagnostic performance of the WHO criteria as a basis and another test for mortality by age as a competing risk. The potential interactions between the study factors were also explored. Stata software version 14 and Microsoft Excel 2013 were used for analysis.

## Results

The sample frame consisted of 7050 individuals. Since a significant loss of data from the medical records was evidenced during the collection of the information, it was decided to collect a sample at least twice the expected number, obtaining a sample of 349 individuals, 133 cases, and 216 controls. The selection process is illustrated in Figure 1.

**Figure 1.**
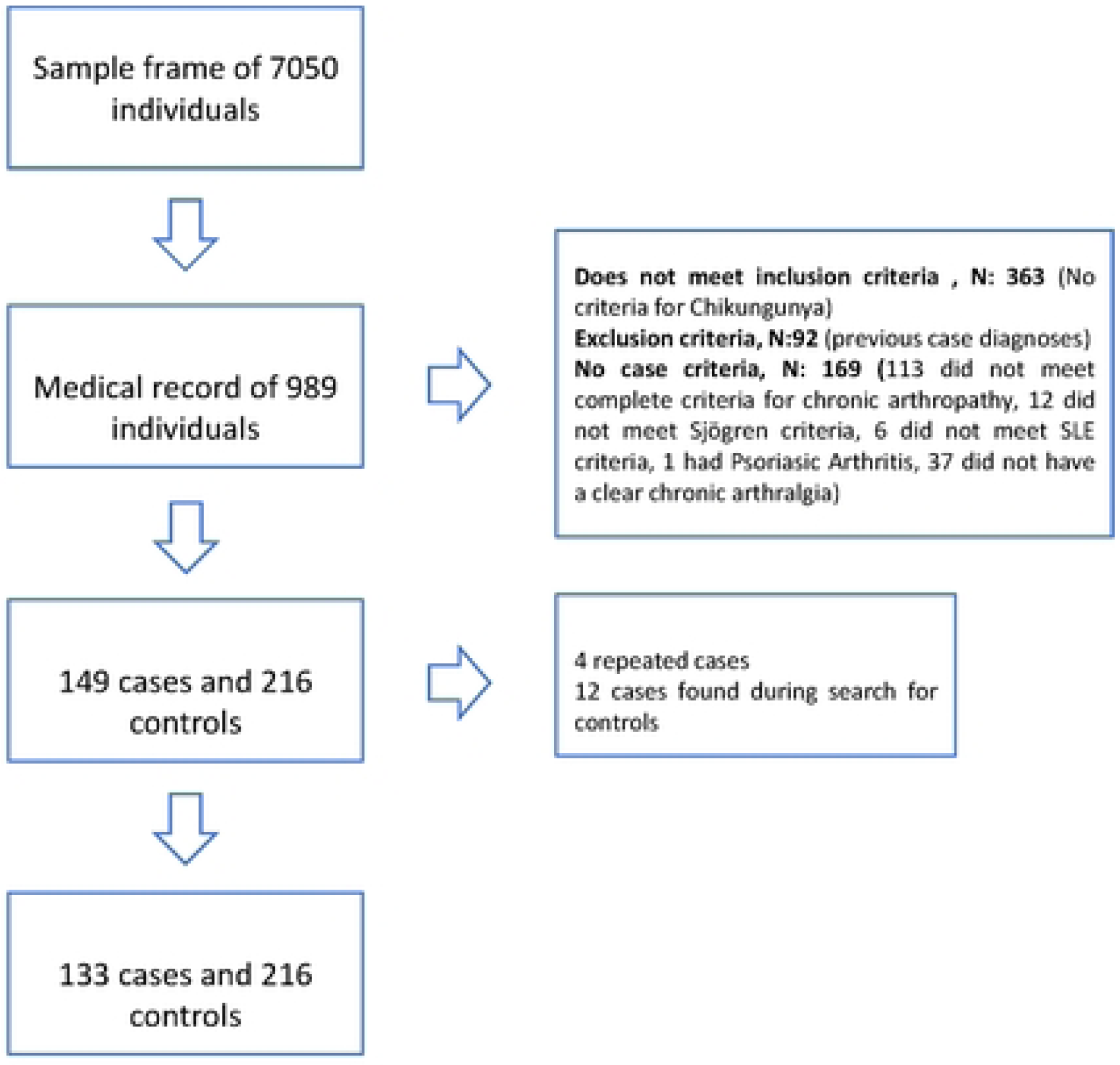
Flowchart for the selection of cases and controls of the study

The first individual with chikungunya in the study dates on January 13, 2015, and the last on December 26 of the same year. Only 1.72% of the individuals in the study achieved a professional degree, and none had a master’s, doctorate, or postgraduate degree. Regarding the behavior of the laboratory variables, a record of C-reactive protein or erythrocyte sedimentation rate was not obtained in a sufficient number of individuals to perform a reliable analysis. No cases of inflammatory spondyloarthropathy were observed in this study.

The cumulative incidence of chronic chikungunya from 2015 to 2019 was 23%, for rheumatoid arthritis was 25.8/10,000 vs. 7.5/10,000 in the general population (p<0.001), for systemic lupus erythematosus was 27/100,000 vs. 25/100,000 (p = 0.78), for Sjögren syndrome 14/100,000 vs. 29/100,000 (p = 0.022), and rheumatic polymyalgia 9.3/10,000 vs. 32/10,000 in the population older than 50 years (p < 0.001). Only Rheumatoid Arthritis had a higher cumulative incidence than the general population.

In the bivariate analysis, *female gender, diabetes, osteoarthritis, COPD* and *duration of arthralgia* were risk factors. In contrast, the involvement of wrists and arthralgia (vs arthritis) were protective factors; the results are presented in Tables 1 and Table A2 (see Supplemental Digital Content 3), respectively. Case subtypes were distributed as follows: one hundred and eleven individuals with chikungunya arthropathy (83,4%), eight with seronegative rheumatoid arthritis (6,02%), seven with seropositive rheumatoid arthritis - positive rheumatoid factor (5,26%), three with seropositive rheumatoid arthritis - positive rheumatoid factor and anti-citrullinated protein antibodies (2,26%), two individuals with systemic lupus erythematosus (1,5%), one individual with Sjögren syndrome (0,75%), and one individual with polymyalgia rheumatica (0,75%).

When the three regression models were made a) with the sociodemographic variables and background of the individual, b) with the clinical variables of the infection, and c) with the laboratory variables, it was found that *female gender*, *osteoarthritis*, and *duration of arthralgia* remained significant after adjustment (p < 0,05). No statistical significance was found in the model that evaluated the laboratories (leukocytes, hemoglobin, and platelets) (data no shown).

Given that there was a loss of data with the model composed of the significant variables from the 3 models previously described (see statistic analysis section), the alternative strategy was carried out in which two models were built: one with the imputed data of the variables with p < 0,2 coming from the bivariate analysis, and the other excluding the variables that had more than 50% of missing data (table A3, Supplemental Digital Content 4).

In both models, the female gender showed evidence of being a risk factor (OR: 2.27, CI95: 1.03 - 4.97, = 0.04 in the imputed data, and OR of 2.37, CI95: 1.09 - 5.17, p = 0.029 in the model that excludes missing data) while acute involvement of the wrists behaved as a protective factor (OR: 0.44, CI95: 0.20 - 0.95, p < 0.037 in the imputed data model and OR of 0.29, 95 CI: 0.13 - 0.65, p = 0.003 in the model that excludes missing data). In contrast, Arthralgia duration was a significant risk factor (OR: 1.03, CI95: 1.00 - 1.06, p = 0.02) just in the imputed data model. In the model that excluded variables with high data losses, acute involvement of the elbows (OR: 2.66, CI95: 1.22 - 5.77, p = 0.013) and the High body mass index (overweight or obesity, OR: 2.1, CI95: 1.00 - 4.42, p = 0.05) were risk factors. The results of both models are presented in Tables 2 and 3.

### Interactions

When evaluating female sex and having diabetes, it was found that although there was a loss of statistical significance of these variables in the model, a statistically significant positive interaction was found between them (p = 0.029). Female sex and having a low educational level showed a positive interaction (p = 0.003), as did being 40 years old or older and overweight or obese, (p: 0.02), age 40 years or older and having a low educational level (p: 0.003), and being diabetic and having a low educational level (p = 0.05) (See figure 2). All variables lost statistical significance in each model. The fact that patients with rheumatological diseases have a higher cardiovascular risk, that cardiovascular events have inflammatory components, and that obesity and smoking are factors associated with rheumatoid arthritis, interactions between dyslipidemia, smoking, diabetes, and hypertension were explored. An interaction was found only between dyslipidemia and hypertension (p = 0.014), without statistical significance for each of the variables in the model.

**Figure 2.**
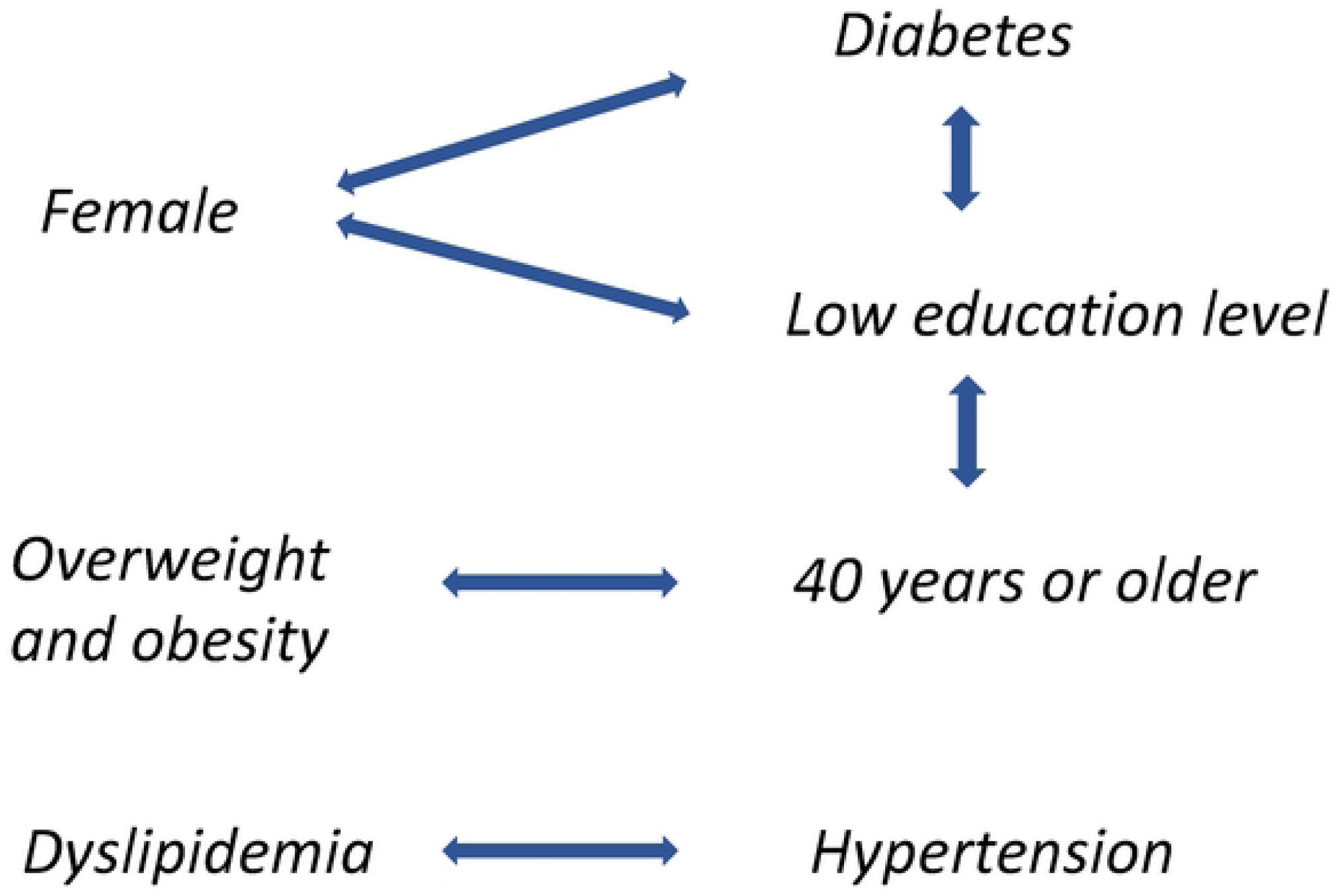
Interactions between variables that increased the probability of developing chronic arthropathy or rheumatological disease after clinical chikungunya infection.

For the sensitivity analysis of the diagnostic inaccuracy of the WHO criteria for chikungunya, a probability of 7% of potential false positives in the sample was calculated (see supplemental digital content 5). Based on the low incidence of rheumatic diseases in the general population, an initial analysis was performed assuming that the false positives were most likely controls and that the distribution of the exposition variables in these was similar to that of the study sample in the case of sex and age, and to that of the general population for variables for which data were available (28–32). Only COPD showed an increase in the magnitude of the association.

A second analysis was performed assuming extreme behavior of the data where all false positives were grouped each time in a quadrant of the 2 x 2 table for cases and controls. The results were highly variable and many of them were conditioned by a low number of individuals in some cells (data not shown). Although these scenarios are assumed to be unlikely, they can be considered for the analysis of the studies about behavior of a new epidemic when it does not develop as expected. Finally, it was calculated that at least six individuals in the control group over 40 years of age could have died. Assuming that they could have been cases, the ORs for age were recalculated and in the bivariate analysis the OR went from 1.49 (95CI: 0.93 - 2.39, p = 0.07) to 1.69 (95CI: 1.06 - 2.7, p = 0.01) raising the possibility of age as an unadjusted risk factor.

## Discussion

This study identified factors associated with the development of chronic arthropathy and/or rheumatic diseases in a specific population of Valle del Cauca, including other findings that should be mentioned. The majority of individuals affected by symptomatic chikungunya were women; it is unclear whether this represents a greater susceptibility to infection or a greater frequency of consultation with health services. On the other hand, there was little representation of high educational levels that somehow reflected the Colombian population’s distribution of education, where only a few reached professional and postgraduate levels.

When the frequencies of rheumatoid arthritis, systemic lupus erythematosus, Sjögren’s, and polymyalgia rheumatica were evaluated in the sample frame, a deficient representation of these conditions was found, except for rheumatoid arthritis, which was also the only one that presented an annual incidence higher than that expected in the general population. This suggests that the publications or reports that have described individuals with systemic lupus erythematosus, Sjögren syndrome, ankylosing spondylitis, and polymyalgia rheumatica after chikungunya may show only fortuitous associations. When performing all the calculations of the study but excluding the individuals with these diagnoses (four in total, corresponding to 1.14% of the sample), there was no significant change in the results (data not shown). In the case of chronic chikungunya, the prevalence was lower than that reported in other studies, which reached up to 40% (33). This is possible because these studies were based on self-reports obtained through interviews and included a wide range of symptoms, not only in joints but also in muscles and tendons. However, the underreporting of symptomatic patients who did not attend health services cannot be ruled out in this study.

Regarding the main objective of the study, it should be mentioned that the female sex was one of the factors associated with the development of rheumatological disease or chronic arthropathy, which is consistent with other studies, and with the fact that rheumatological conditions are more frequent in women than in men, except for spondyloarthropathy, which was not found in the present study (17,23,26). As inflammatory arthropathies are diseases with complex features, it is possible that chikungunya interacts with the peculiarities inherent to the female sex, such as hormonal, social, and environmental factors, and triggers the development of the disease (34,35).

By what happens in arthritis and polymyalgia rheumatica, the age in the crude analyses behaved as a risk factor. It could be that the senescence of the immune system explains it because of its loss of regulation and tolerance over time (35); however, the explanation is necessarily more complex and it can be related to the multiple biological and social changes strongly associated with the life cycle of the individuals that, in the present study and other similar ones, can explain the loss of statistical significance when adjusting for other variables.

Osteoarthritis is another factor found in the crude analysis and is similar to previous studies. It is difficult to conclude in this type of study that this condition is predisposing because although it is plausible that joint structural alterations increase susceptibility to inflammatory phenomena, memory biases can generate false associations, or, as in the present study, be confused by other variables, especially age.

The duration of arthralgia in the acute stage was a consistent risk factor in this and other studies. Despite being one of the variables with the most missing data, it behaved as a risk factor in the crude analysis, the regression model that only evaluated the characteristics of the infection, and the model with imputed data. It may be the manifestation of an individual’s predisposition for the symptom to remain chronic, but it can also respond to the presence of a gradient in which the more significant the intensity and/or initial duration of the symptoms, the greater the probability that it will become chronic, as other studies have suggested, where pain intensity, the persistence of viral load over time, absenteeism from work, and high levels of IL-6 and immunoglobulin G are risk factors for chronic chikungunya arthropathy (5,7,18,19,21,22,25)

Regarding the involvement of the elbows as a risk factor and the wrists as a protective factor, although it may be just chance and/or confounded by other factors, different rheumatological pathologies have a propensity for certain types of joints. For example, rheumatoid arthritis is located predominantly in the hands and small joints symmetrically; polymyalgia rheumatica compromises the bursae of the pelvic and shoulder girdle, while spondyloarthropathies are located in large joints asymmetrically. Thus, joint location may respond to the particularities of individuals that predispose or protect them against the development of chronic arthropathy after chikungunya infection. Finally, the finding of arthralgia as a protective factor when compared to arthritis is reciprocal to the fact that arthritis behaves as a risk factor, which coincides with the possible existence of a severity gradient as previously described.

The interactions found in this study suggest that intervening conventional risk factors such as overweight, diabetes, hypertension, or dyslipidemia, and in social determinants of health, such as the educational level in this case, could reduce the risk of developing chronic joint pathology in the event of a new outbreak of chikungunya.

This study has several limitations. First, it was not possible to select the individuals with laboratory tests because the source of information was medical records. In exchange, the medical criterion was appealed to, which is considered a good approximation if one believes that professional training and expertise better classify patients and their clinical symptoms when compared to self-report or telephone interviews.

Because of the limitation in the number of individuals other than mestizo ethnicity, it was not possible to make inferences related to this factor, extending the same restriction to the area of residence since the vast majority belong to the urban area. This reflects the type of individuals in the study, including subjects insured by the Colombian health system in the contributory regime (salaried workers and their families). Similarly, the low representativeness of the extreme socioeconomic strata and high educational levels limit the extrapolation of the results to the entire population. In any case, this study represents one of the most important groups of the Colombian population (mestizo, salaried individuals living in urban areas) for whom the information in the study is helpful, including the development of specific public policies.

Important information, such as socioeconomic status, was not available, although details on educational level were obtained, which provides similar information. Nevertheless, it is essential to obtain this information in future studies.

Nine variables had more than 50% data loss (23% of the variables). The sample size which was almost three times higher than the calculated one, and the imputation of the data, compensated for this limitation.

Finally, information based on medical records was limited to those who attended health services. For this reason, there is the possibility of a selection bias towards sicker individuals, with better socioeconomic status, and female predominance. If low socioeconomic status and then a low educational level are risk factors for case development, this association may have been diluted.

Despite the limitations described, the study answered the main question and specific objectives adequately and was able to explore the relationship between different rheumatological diseases and chikungunya infection.

## Conclusions

The present study found that female gender and the duration of joint symptoms in the acute phase were the main risk factors for the development of chronic chikungunya arthropathy or rheumatological pathology in Valle del Cauca, Colombia. Wrist involvement was found to behave as a protective factor. In this study, only rheumatoid arthritis was found to be a rheumatological disease associated with chikungunya infection. Finally, the study suggests that conventional cardiovascular risk factors such as hypertension, diabetes, obesity, dyslipidemia, and low educational level, could interact with each other and enhance the probability of developing chronic symptoms after clinical infection by the chikungunya virus.

A prospective, population-based study with a laboratory diagnosis of infection is necessary to resolve the limitations and clarify the questions that arose during the development of this research.

## Data Availability

All relevant data are within the manuscript and its Supporting Information files.

## Take-home messages

- Chikungunya infection can cause chronic joint pain in up to 30% of the affected patients.
- Female gender and prolonged duration of joint symptoms in the acute phase are risk factors for developing chronic symptoms, whereas wrist involvement is likely to be a protective factor.
- The simultaneous presence of hypertension, diabetes, obesity, dyslipidemia, and low educational level in an individual seems to increase the risk of chronic joint symptoms after chikungunya infection; therefore, intervention may be a strategy to reduce the risk of this complication.
- Rheumatoid arthritis appears to be the only rheumatological disease associated with chikungunya infection. Further studies are required to clarify this association.

## Notes

### Competing Interest Statement

The authors have declared no competing interest.

### Funding Statement

The author(s) received no specific funding for this work.

### Author Declarations

The study was approved by the Ethics Committee of the Faculty of Health of the Universidad del Valle Colombia. CIREH and the Ethics Committee of the HPI from which the study subjects came (approved record 007-93 020).

